# Blood lactate kinetics as biomarkers of MRI brain injury in neonates with hypoxic-ischemic encephalopathy

**DOI:** 10.1101/2025.09.19.25336141

**Authors:** Giulia Bassani, Marion Decaillet, Patric Hagmann, Jean-Baptiste Ledoux, Anita C. Truttmann, Juliane Schneider

## Abstract

**Background:** In neonates with hypoxic-ischemic encephalopathy (HIE), early biomarkers are needed to enhance prognostic accuracy. We hypothesized that blood lactate kinetics correlate with magnetic resonance imaging (MRI)–based brain injury severity.

**Methods:** In this prospective cohort study, neonates with HIE admitted to a tertiary neonatal intensive care unit underwent brain MRI within the first week of life. Injury severity was graded using a validated MRI scoring system, with neonates categorized into low severity (LS) or high severity (HS) groups based on the 75th percentile of the total score. Lactate kinetics were evaluated through peak lactate levels, time to lactate normalization (TLN), and area under the curve (AUC). Associations between lactate kinetics and MRI scores were analyzed.

**Results:** Among forty-eight neonates, 83% underwent therapeutic hypothermia. Compared with the LS group, the HS group had more seizures, higher Thompson and Sarnat scores, and more abnormal neurological exams at discharge. Peak lactate was higher in the HS group (*p*=0.02) and correlated with MRI grey matter subscores (*p*=0.004). Lactate AUC and TLN were positively associated with MRI total and grey matter scores (all *p*<0.01).

**Conclusion:** Lactate kinetics are associated with MRI-assessed brain injury severity in HIE and may help stratification to tailor therapeutic strategies.

**Impact:**

- Blood lactate kinetics are associated with MRI-assessed brain injury severity in neonates with HIE.
- This study highlights the value of serial lactate measures, beyond single time-point levels, in early prognostication.
- Combining biochemical markers with imaging scores may improve the identification of infants at high risk of adverse outcomes.
- These findings support the potential use of lactate kinetics to stratify patients and guide future therapeutic strategies.

## BACKGROUND

Hypoxic-ischemic encephalopathy (HIE) is a clinical syndrome of neurological dysfunction caused by a perinatal event of diminished blood and oxygen supply. In high-income countries HIE is estimated to affect between 0.5 and 1.5/1000 live births^1^ and remains a major cause of death and lifelong disability worldwide.^2^ Survivors may develop motor deficits, functional disabilities and cognitive impairments later in childhood.^3^ Therapeutic hypothermia (TH) is the only treatment proven to safely improve both mortality and neurodevelopmental outcomes, with a number needed to treat of 6.^4^ Several adjuvant therapies are under investigation, but none has yet demonstrated their effectiveness.^5^ Clinical scoring systems, such as the Sarnat^6^ and Thompson^7^ scores, are used to assess the severity of encephalopathy, yet their prognostic accuracy alone is not sufficient.

To improve prognostication and guide individualized care, there is a growing interest in identifying early biomarkers of brain injury in HIE. Various biochemical and physiological markers have been proposed, including S100B, neuron-specific enolase, glial fibrillary acidic protein, and circulating microRNAs, but none have reached widespread clinical use.^8, 9^ In clinical practice, pH and base deficit from cord or early neonatal blood remain the most common indicators,^10^ reflecting metabolic acidosis in HIE. Cord lactate, strongly correlated with pH and base deficit,^11^ has thus been proposed as an additional marker of impaired metabolism in HIE. In addition to protein biomarkers, metabolomic approaches have also been investigated to better characterize the biochemical alterations associated with HIE, revealing changes in key metabolites such as lactate, pyruvate, tricarboxylic acid, cycle intermediates, and ketone bodies, which reflect profound disruption of energy metabolism at birth.^12, 13^ Among the metabolites most consistently altered in HIE, blood lactate has emerged as a relevant biomarker, as elevated or sustained levels have been repeatedly associated with encephalopathy severity^14–19^ and others adverse outcomes as seizure burden.^20^ However, most of these studies have relied on isolated measurements rather than analyzing lactate kinetics, which may offer a more dynamic and informative assessment of metabolic recovery.

While metabolic biomarkers are promising, current prognostic assessment in HIE still relies heavily on brain magnetic resonance imaging (MRI). MRI is the imaging modality of choice for detecting injury patterns associated with poor outcomes.^21–24^ Several validated MRI scoring systems have been developed to assess the severity of brain injury in HIE, most were designed to be applied to T1- and T2-weighted sequences only.^25, 26^ As such, they typically rely on imaging performed in the second week of life, since abnormalities on these conventional sequences may still be subtle or absent during the first days following the hypoxic-ischemic insult.^27^ In contrast, the scoring system recently proposed by Weeke et al.^28^ incorporates diffusion-weighted imaging (DWI), allowing it to be applied to earlier MRI scans. This comprehensive score evaluates lesions across the grey matter (GM), white matter (WM), and cerebellum, and has demonstrated strong predictive value for neurodevelopmental outcomes both at 2 years and at school age.^28^ Furthermore, in a recent comparative analysis, Weeke’s scoring system showed superior predictive performance for adverse neurodevelopmental outcomes compared with the historical Barkovich score.^29^

Our prospective cohort study aimed to investigate the association between blood lactate kinetics, including peak value, time to lactate normalization (TLN), and area under the curve (AUC), and the severity of brain lesions, as graded by the MRI scoring system developed by Weeke et al.^28^ As an easily obtainable and routinely measured biomarker, lactate could help refine early prognostic accuracy and support the stratification of neonates with HIE who may benefit from targeted therapies or further investigations.

We hypothesized that a higher peak lactate level, a longer TLN, and a greater AUC would be positively associated with increased injury severity on MRI.

## METHODS

### Study design and setting

This is a single-center prospective observational cohort study conducted at the level III Neonatal Intensive Care Unit of the Lausanne University Hospital, Switzerland. The study was approved by the Ethics Committee of the Canton of Vaud (Commission cantonale d’éthique de la recherche sur l’être humain, CER-VD (project ID 2021-00808). All methods were carried out in accordance with the STROBE 2007 guidelines and with the principle of the Declaration of Helsinki. Written informed consent was obtained from the parents of all enrolled neonates prior to participation.

### Participants

All term and near-term neonates (≥ 35 0/7 weeks of gestational age), born between February 2022 and December 2024, with all HIE grades, treated or not with TH, were eligible for enrolment in the study.

The diagnosis of HIE was established in neonates presenting clinical signs of neonatal encephalopathy, as described by Sarnat^6^ and meeting criteria for perinatal asphyxia, including: evidence of fetal distress (acute perinatal event, arterial cord pH <7.0 or cord base deficit <16 mmol/L) and evidence of neonatal distress (Apgar score <5 at 10 minutes of life, blood pH <7.0, base deficit <16 mmol/L, blood lactate >12 mmol/L within the first hour of life or prolonged resuscitation requiring bag-mask ventilation for >10 minutes). The clinical severity of HIE was graded as mild, moderate or severe according to the Sarnat scoring system. Exclusion criteria were major malformations, known or suspected metabolic, cardiac, or genetic abnormalities, refusal of consent or inability to obtain it.

### Protocol

Whole-body TH (target core temperature: 33–34°C) was initiated within the first 6 hours of life in neonates fulfilling the criteria, i.e. evidence of fetal distress and neonatal distress as described above, presence of HIE Sarnat stage > I or Thompson score ≥ 7 and abnormal continuous amplitude integrated EEG (aEEG) background. TH was maintained for 72 hours followed by rewarming at a rate of 0.2-0.3° C/h until a core temperature of 36.5°C was reached.

All neonates underwent routine standard-of-care neuromonitoring, including continuous aEEG for 96 hours, one-hour EEG recording during the first 24 hours of TH and at rewarming, regular neurological examinations, head ultrasound and brain MRI.

### Measurements

#### Lactate kinetics

Blood gases (capillary, venous or arterial, mostly arterial) were measured as clinically indicated, usually every 1 to 4 hours. We collected blood lactate values until the patient discharge or death. TLN was defined as the time from birth, in hours, until the lactate level was ≤ 2.5 mmol/L. Maximal blood lactate was defined as the higher value collected until patient discharge or death. The AUC of lactate levels over time was calculated to assess their temporal evolution during the study period. We computed the AUC using trapezoidal integration of lactate measurements, processed with the *pracma*package (v1.9.9) via its *trapz()* function. The results are expressed in (mmol*h/L).

#### Demographic and clinical data

Other clinical variables were prospectively collected including resuscitation needs, Apgar score, Sarnat stage, Thompson score, ventilation requirements (duration of invasive/non-invasive ventilation, highest mean airway pressure), sepsis, presence of clinically suspected seizures, and additional blood gas parameters (base excess, pCO2, bicarbonate). Glycemia, liver function, creatinine level, and coagulation tests were also routinely measured and recorded. Demographic data, as well as fetal and delivery characteristics, were also documented (see Table 1). A neonatologist experienced in neurodevelopment performed a detailed neurological examination before discharge.

**Table 1.**
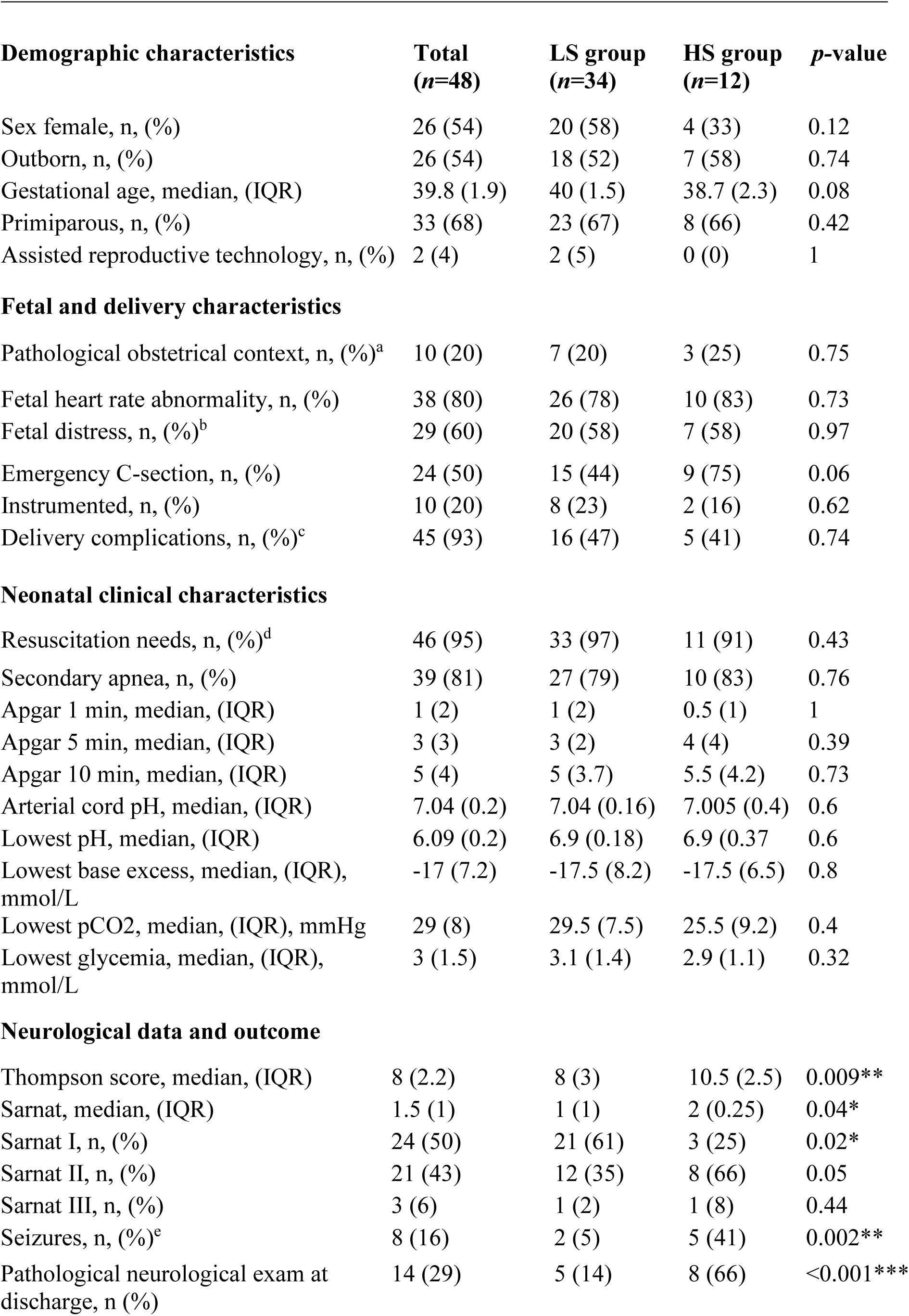
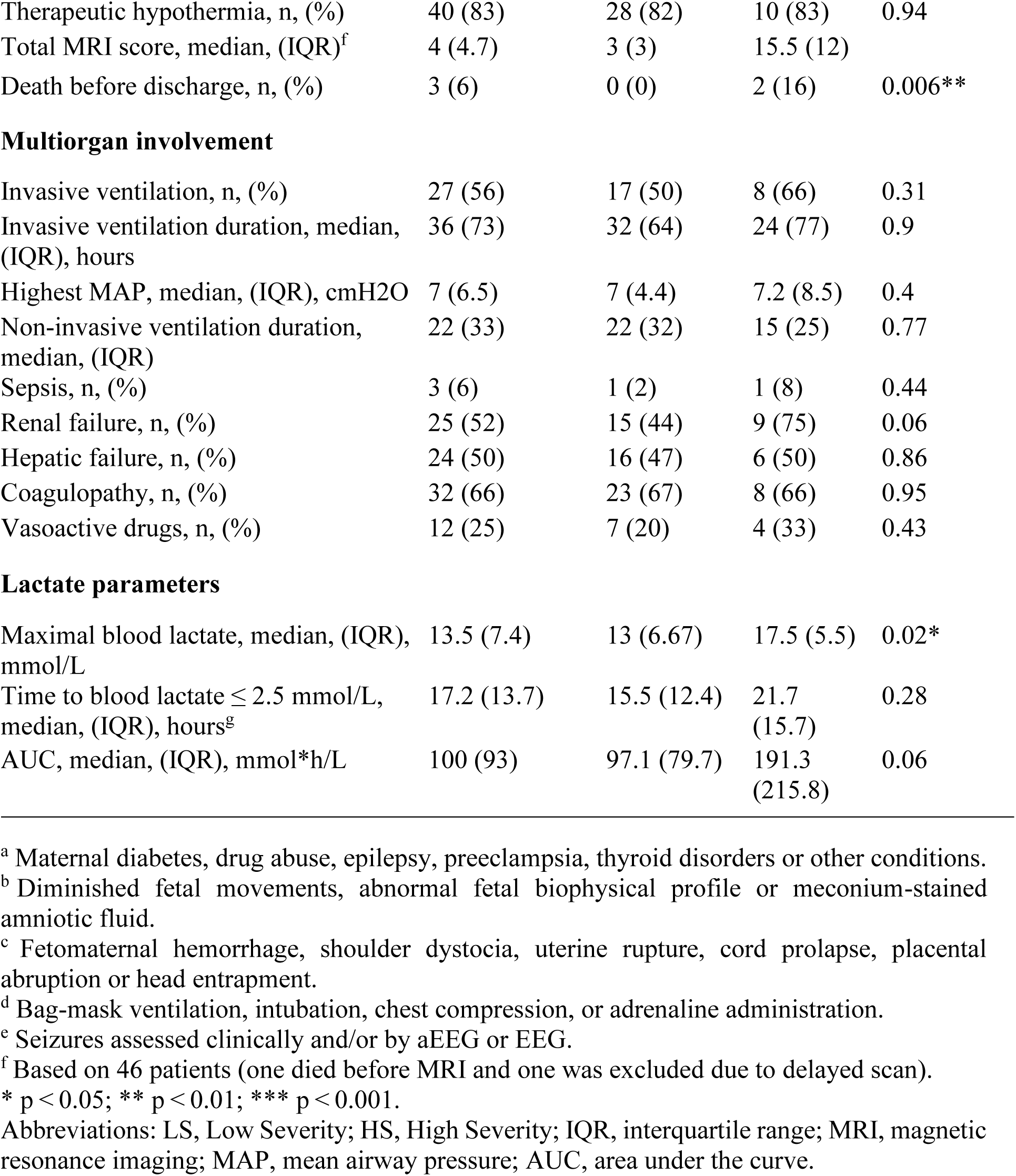
Demographic and clinical data by MRI severity group.

#### Brain MRI

Brain MRI was performed between the 4th and 7th day of life, and after rewarming if infant received TH, using a 3-Tesla PrismaFit system (Siemens Healthineers®; Forchheim, Germany), in natural sleep or mild sedation if necessary (chloral hydrate, 20 mg/kg enterally; hospital pharmacy, CHUV, Lausanne, Switzerland). Neonates were transported in an MRI-compatible incubator equipped with an integrated 16-channel head coil (Nomag® LMT Medical Systems, Lübeck, Germany), which remained in place during the scan. Acoustic ear protection (MiniMuffs®, Natus Medical, San Carlos, CA, USA) was used to reduce scanner noise and promote sleep. Support foams were also applied to stabilize the head position and prevent motion artifacts during the scan. Monitoring of temperature, heart rate, and oxygen saturation was provided. A nurse and a physician were present as needed throughout the procedure.

The MR protocol included the following sequences (see Supplementary Table): 1) Sagittal T1-MP2RAGE (Magnetization Prepared 2 Rapid Acquisition Gradient Echo) with a 1mm^3^ isotropic voxel size. 2) Axial T2-weighted turbo spin echo with a 0.4x0.4x1.6mm^3^ interpolated voxel size. 3) Coronal T2-weighted turbo spin echo with a 0.4x0.4x1.2mm^3^ interpolated voxel size. 4) Axial T2-weighted gradient echo with a 0.6x0.6x3mm^3^ voxel size. 5) Axial DTI (diffusion tensor imaging) with a 1.7mm^3^ isotropic voxel size.

A neuroradiologist reviewed all the images, determining the nature, location and severity of the lesions. In parallel, a neonatologist trained in neonatal neurology and a 6-year experienced neonatologist assessed independently all MRI scans using the score described by Weeke et al.^28^ In case of disagreement, consensus was obtained through joint review of the images. The scoring was compared with the neuroradiologist’s report for further verification. The applied score assesses brain injury in 3 areas, giving distinct subscores: deep GM, cerebral WM/cortex, and cerebellum. A fourth subscore (additional) assesses the presence of intraventricular or subdural hemorrhages and sinovenous thrombosis. The total score was calculated by adding the 4 subscores (GM, WM, cerebellum, additional), yielding a maximum possible score of 55.

The primary outcome was the correlation between lactate kinetics (maximal lactate level, TLN, AUC) and brain injury assessed on MRI. The secondary outcomes were the relationship between lactate kinetics and presence of seizures, treatment with TH or neurological exam at discharge.

### Data analysis

The distribution of the variables, evaluated using the Shapiro-Wilk test, deviated from normality. Consequently, non-parametric tests were used. Descriptive analyses were performed, including the total count and percentage for categorical variables and median with interquartile range (IQR) for continuous variables. Patients were stratified into two groups according to the 75th percentile of the total MRI score (6.7): a low severity (LS) group (score < 6.7), and a high severity (HS) group (score ≥ 6.7).

Differences between the two groups were assessed using the chi-square test (or Fisher’s exact test where appropriated) for categorical data and with the Welch’s test for continuous variables. A linear mixed model was performed to evaluate the difference in changes over time in blood lactate levels between the two groups. Non-parametric Spearman rank-order correlations were computed to assess the relationship between maximal lactate, TLN, lactate AUC, and MRI scores (total score, WM and GM subscores). To account for multiple comparisons, we applied the False Discovery Rate (FDR) correction.

Further analyses were performed based on the correlations results, using General Linear Models (GLM) to assess the relationship between lactate parameters and MRI scores, accounting for confounding variables including sex, gestational age, 5-min Apgar score, Thompson score and presence of clinical seizures.

In addition, the Welch’s test was used to assess differences in lactate parameters (maximal lactate, TLN, lactate AUC) between neonates with or without seizures, those who did or did not receive TH, and those with a normal or pathological neurological exam at discharge.

Analyses were computed with Jamovi (The Jamovi Project, Sydney, Australia; version 2.4.8),^30^ IBM SPSS Statistics (IBM Corp., Armonk, NY, USA; version 29.0.1.0),^31^ and R (R Foundation for Statistical Computing, Vienna, Austria).^32^ *P* values <0.05 were considered significant.

## RESULTS

### Patient characteristics

Among 73 eligible patients, 48 neonates (26 females, 54%) with a median gestational age of 39.8 weeks (IQR 1.9) were enrolled (see Fig. 1 for exclusion reasons). Among the patients who were enrolled, 3 (6%) died during the neonatal period. Causes of death included withdrawal of care due to anticipated poor neurological outcome (n=2) and death despite full intensive support (n=1). Two patients were excluded from the correlation analysis: one who died before undergoing MRI, and another whose MRI was performed on day 10 of life.

**Figure 1.**
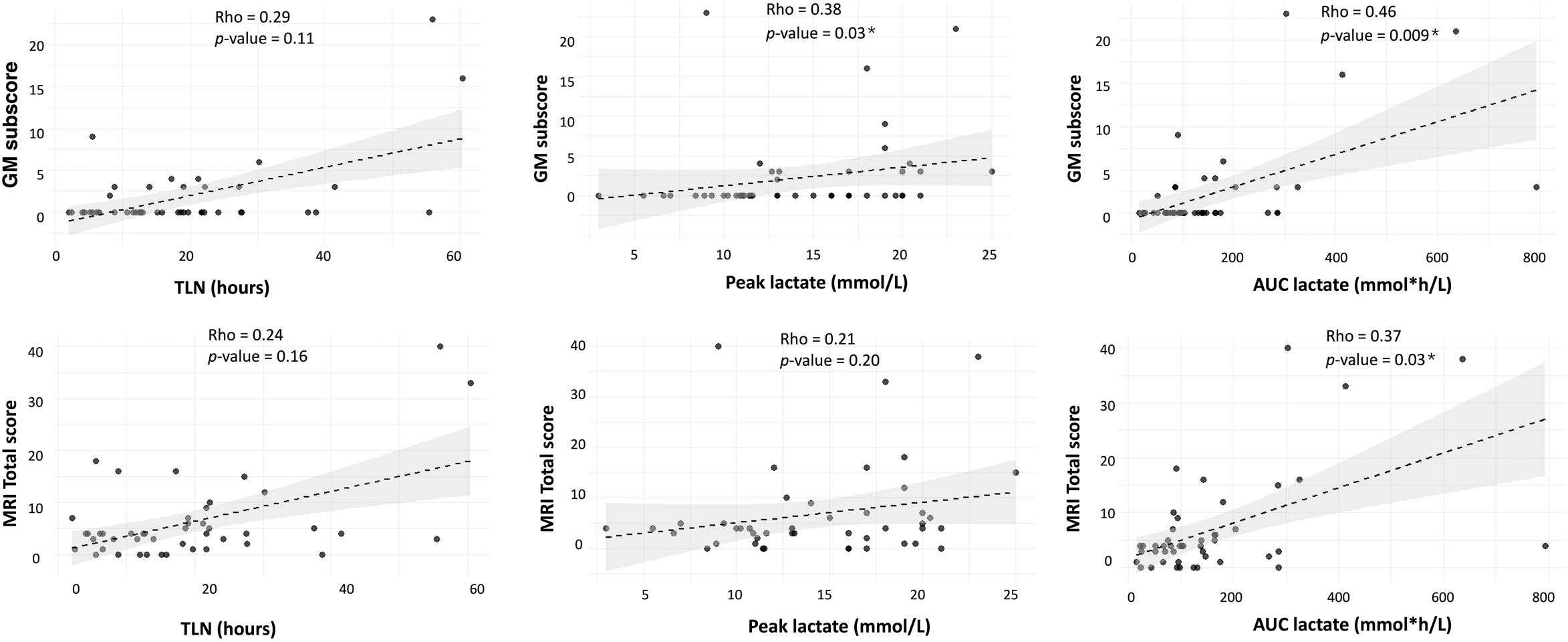
Flowchart of the study population. Patients enrolled and undergoing early MRI were classified into low or high severity groups based on the 75^th^ percentile of the total MRI score. Abbreviations: MRI, magnetic resonance imaging; DOL, day of life; LS, low severity; HS, high severity.

Based on the admission Sarnat examination, 24 (50%) neonates presented with mild, 21 (43%) with moderate and 3 (6%) with severe encephalopathy. Forty (83%) patients underwent TH. Among the 46 neonates with interpretable MRI data, 34 (74%) had a LS injury score and 12 (26%) a HS score, as assessed using the MRI-based severity scoring system. In the entire cohort, the total MRI score ranged from 0 to 40, with a median of 4 (IQR 4.7). The median total MRI score was 3 (IQR 3) in the LS group and 15.5 (IQR 12) in the HS group.

Demographic characteristics, fetal and delivery variables, and clinical parameters were compared between the LS and HS groups (see Table 1). Compared with the LS group, neonates in the HS group had a significantly higher incidence of seizures (*p*=0.003), higher Thompson (*p*=0.009) and Sarnat scores (*p*=0.04), and were more likely to exhibit abnormal neurological findings at discharge (*p*<0.001). Regarding the other variables, no statistically significant differences were observed, not even for Apgar scores or resuscitation measures.

### Primary outcome - lactate kinetics/MRI score correlation

The peak blood lactate was significantly higher in the HS group (median 17.5 mmol/L, IQR 5.5) than in the LS group (median 13 mmol/L, IQR 6.6; *p*=0.02) as illustrated in Fig. 2. Median TLN was 15.5 h (IQR 12.4, n=34) in the LS group and 21.7 h (IQR 15.7, n=11) in the HS group (*p*=0.28). In a random-intercept model, blood lactate levels decreased significantly faster in the LS group compared to the HS group (β = –0.034, p = 0.04). However, when individual variability in the rate of decrease was accounted for in a random slope model, this difference was no longer statistically significant, indicating substantial heterogeneity in lactate trajectories within each group. Blood lactate trajectories over time in both severity groups are shown in Fig. 3. One patient failed to normalize lactate level, with a minimum value of 2.7 mmol/L at 80 h of life, followed by a subsequent increase due to sepsis, which ultimately led to death despite maximal supportive care.

**Figure 2.**
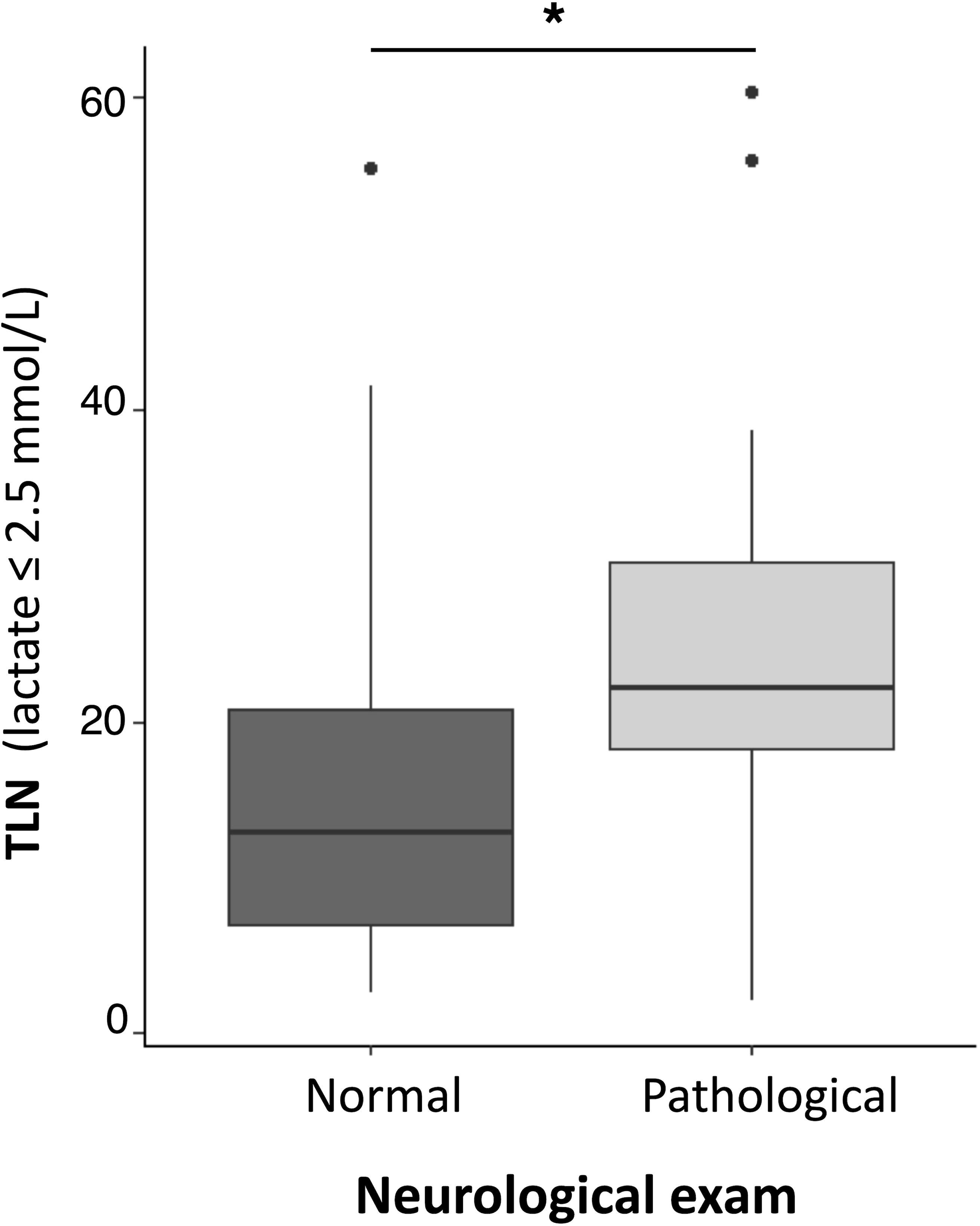
Box plot of peak blood lactate levels in the high-severity (HS) and low-severity (LS) groups. Peak blood lactate was significantly higher in the HS group (median 17.5 mmol/L, IQR 5.5) compared to the LS group (median 13 mmol/L, IQR 6.6; *p* = 0.02).

**Figure 3.**
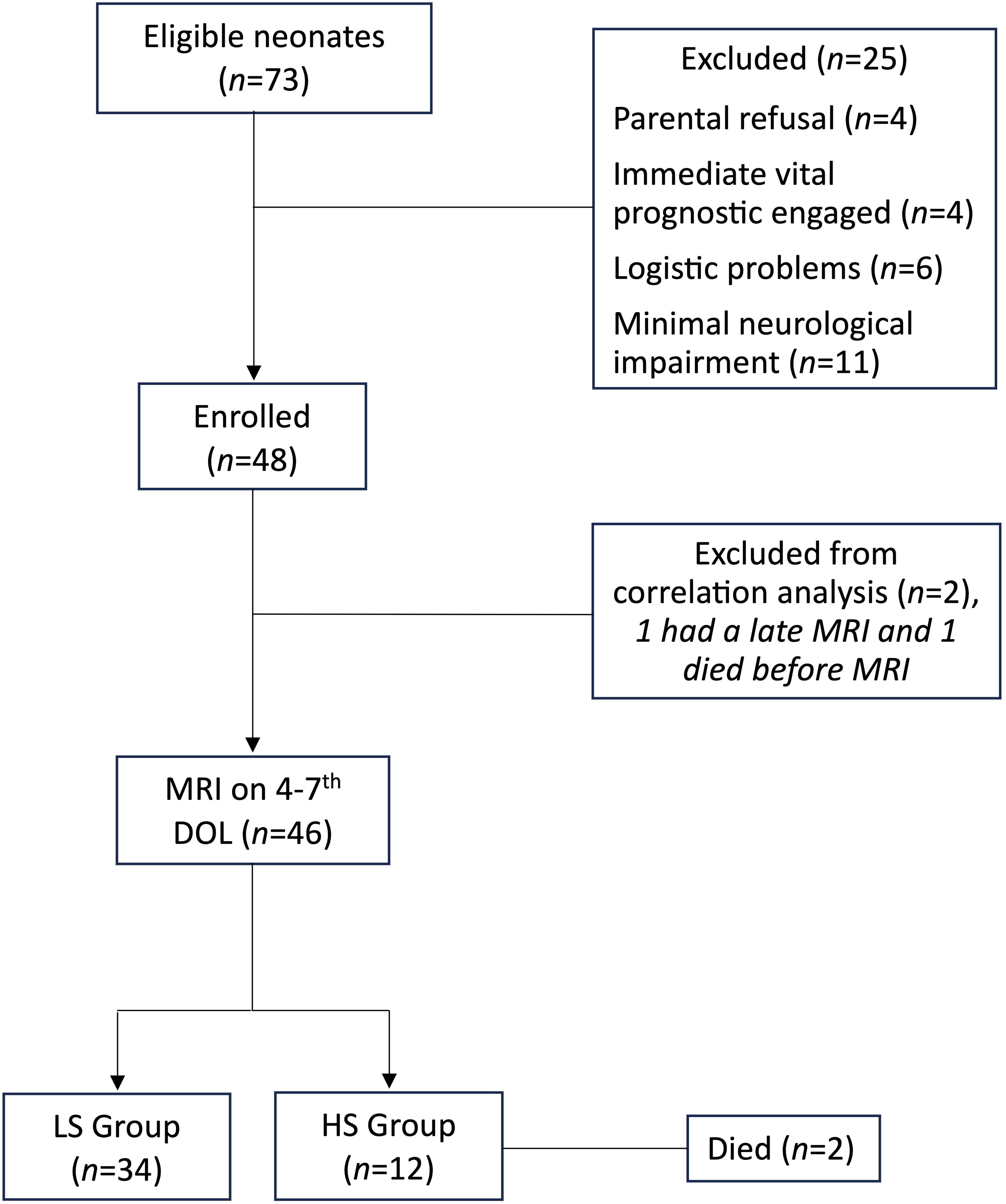
Blood lactate trajectories over time in the low-severity (LS) and high-severity (HS) groups. Each point represents an individual measurement. Random-intercept model: β = –0.034, p = 0.04.

Non-parametric Spearman rank-order correlations with FDR correction revealed that lactate AUC was positively correlated with MRI total score (Rho=0.37, *p*=0.03), as well as with the GM subscore (Rho=0.46, *p*=0.009). Moreover, peak blood lactate was positively correlated with the GM subscore (Rho=0.38, *p*=0.03) (Fig. 4). Other correlations between the MRI scores and lactate kinetics were not statistically significant.

**Figure 4.**
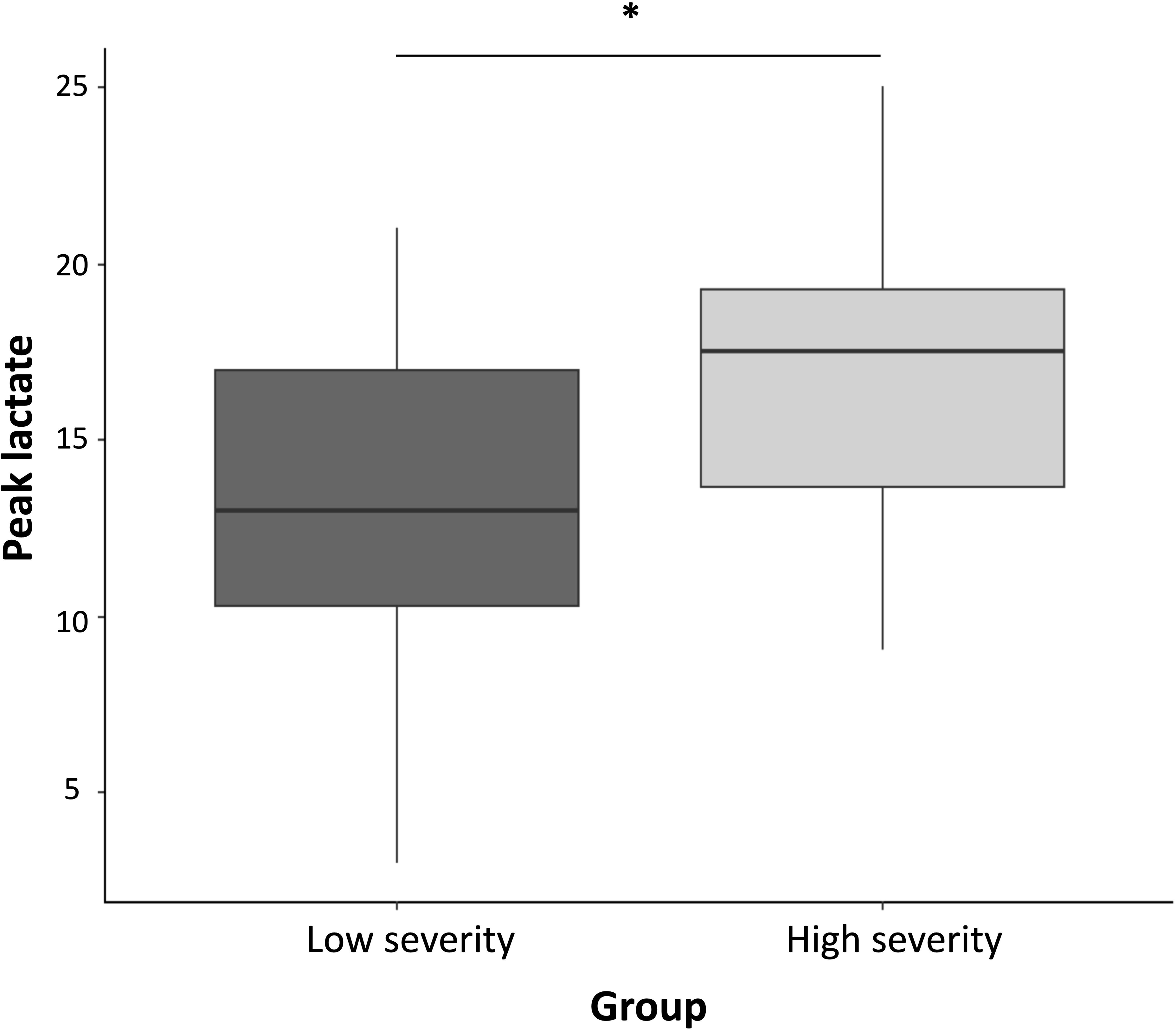
Relationship between TLN, peak lactate, and AUC with MRI total score and grey matter subscore. Shaded areas indicate 95% confidence intervals. * Denotes statistical significance. Abbreviations: TLN, time to lactate normalization (blood lactate ≤ 2.5 mmol/L); AUC, area under the curve; MRI, magnetic resonance imaging.

Complementary analyses using GLMs showed significant positive associations between lactate AUC or TLN and both the total MRI score and the GM subscore (all *p*< 0.01), after adjusting for confounders (i.e., sex, gestational age, Thompson score, 5-min Apgar score and clinical seizures). No associations were found between the peak blood lactate and the MRI scores (see Table 2).

**Table 2.**
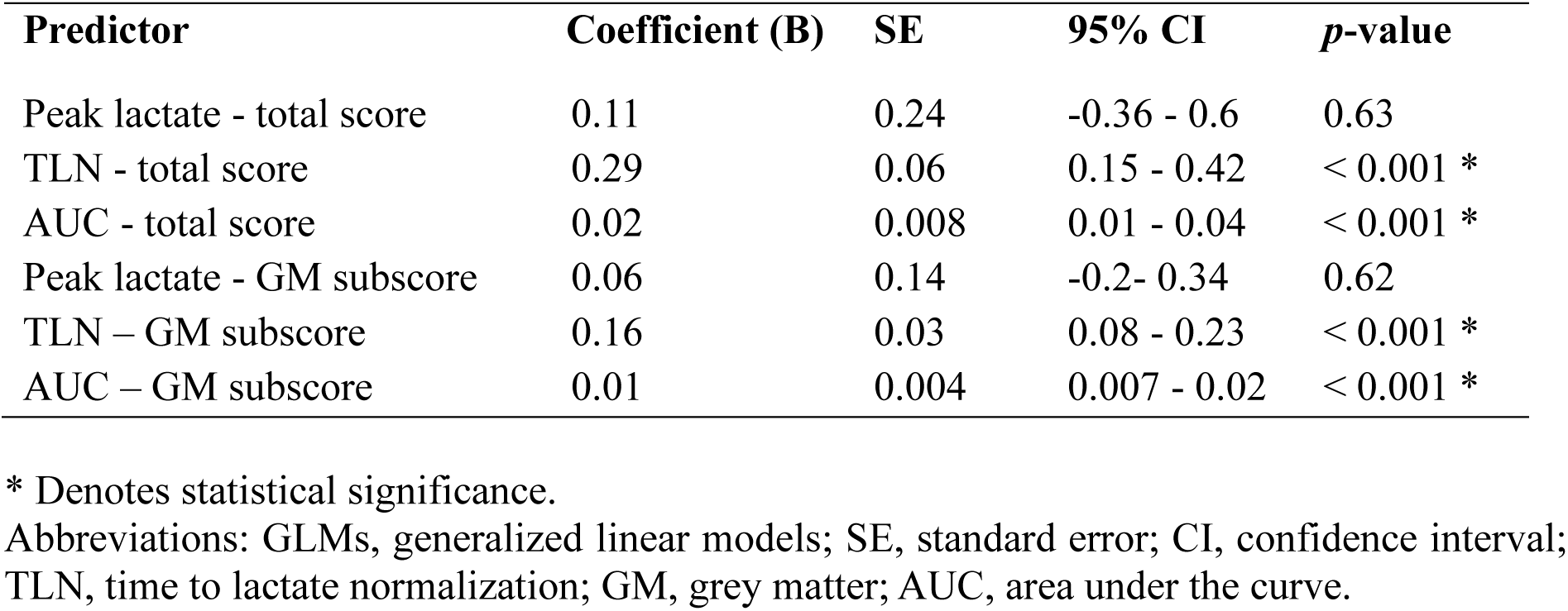
Association between lactate kinetics and MRI injury scores (GLMs).

### Secondary outcome - lactate kinetics/neurological outcome at discharge, seizures and TH

Regarding the secondary outcomes, while TLN was significantly longer in patients with pathological neurological exam at discharge compared to those with a normal neurological exam (median_patho_ = 22.2, IQR_patho_ = 12 vs median_normal_ = 12.8, IQR_normal_ = 13.9, *p*=0.02; Fig. 5), maximal lactate and lactate AUC did not differ between the two groups. Lactate kinetics parameters did not differ significantly between patients with or without seizures, nor between those who received TH and those who did not.

**Figure 5.**
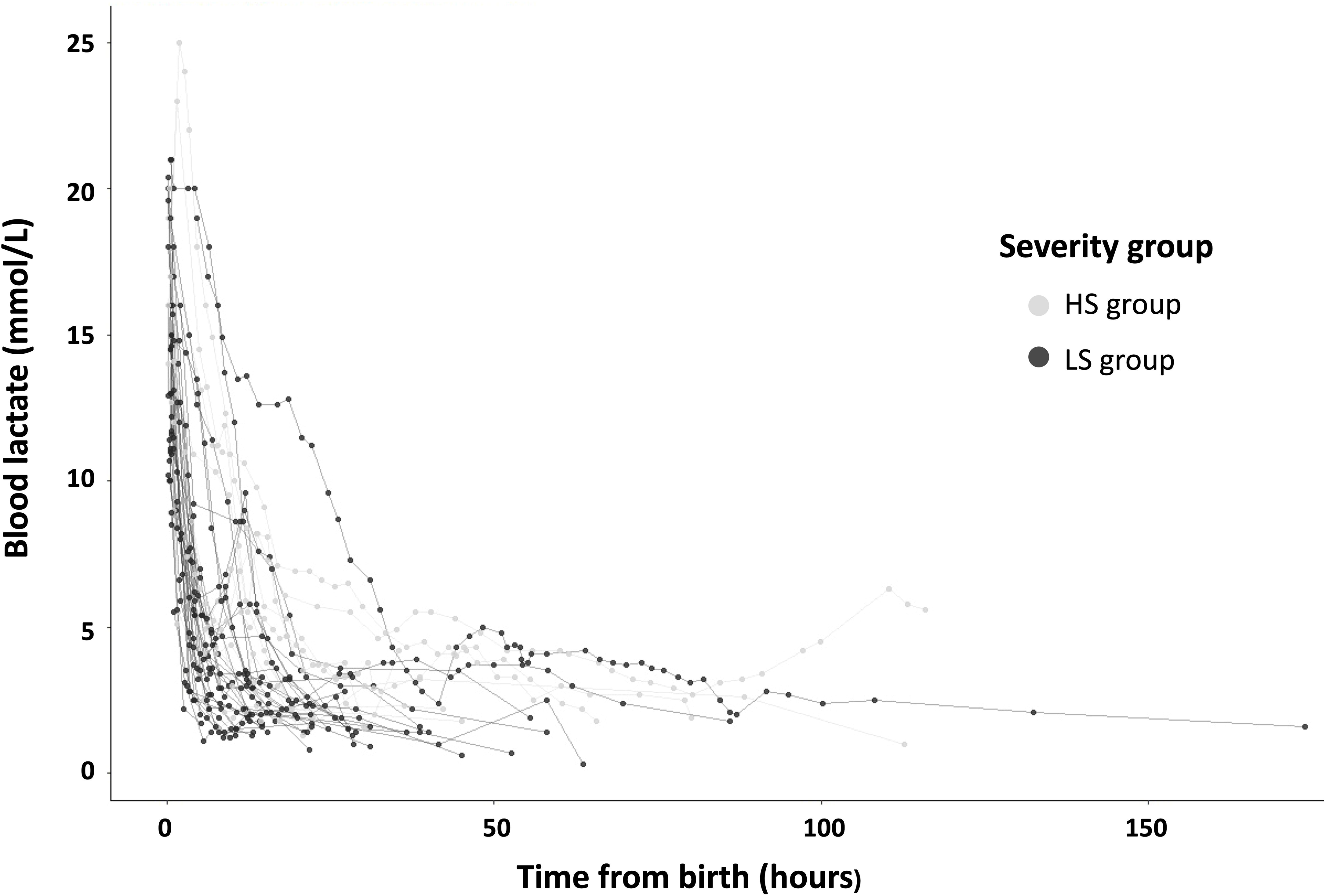
Box plot showing TLN according to neurological exam at discharge. TLN was significantly longer in patients with pathological exam (*p* = 0.02). TLN, time to lactate normalization (blood lactate ≤ 2.5 mmol/L).

## DISCUSSION

The objective of the present study was to assess the association between lactate kinetics and early MRI findings, with the aim of improving prognostic accuracy and supporting the stratification of patients for targeted therapeutic strategies. Most neonates of our cohort presented with a mild to moderate encephalopathy. Compared to neonates with a HS score, those in the LS group exhibited a shorter TLN, a lower peak blood lactate level, and a lower lactate AUC. Correlation and regression analyses revealed a positive independent association between lactate parameters and the severity of brain injury as assessed by MRI. We employed an MRI scoring system that has been validated as a predictor of adverse outcomes, including death or neurological impairment, at both 2 years and school age.^28^ Our findings suggest that lactate kinetics may serve as an early biomarker of brain injury severity and could support the outcome prediction in neonates with HIE. In certain cases, this may help identify subgroups of patients who could benefit from adjunctive therapies.

Lactic acid is the primary end product of anaerobic metabolism, resulting from the fermentation of pyruvate and generating two molecules of adenosine triphosphate. Under conditions of tissue hypoxia, pyruvate accumulates and is subsequently converted into lactate. Thus, circulating lactate is considered as a reliable indicator of tissue hypoxia.^11^ Lactate may also serve as an important energy substrate for the developing brain, particularly during the neonatal period, where monocarboxylate transporters and the astrocyte-neuron lactate shuttle facilitate its utilization in neuronal oxidative metabolism, a mechanism sometimes referred to as “the good lactate”.^33^ Despite its metabolic role, elevated lactate levels are widely used as a marker of cellular stress and impaired perfusion. In adult and pediatric studies, lactate level and lactate clearance have been associated with neurological outcomes after return of circulation following cardiac arrest.^34, 35^ In neonates with asphyxia, several studies have explored the association between lactate levels and lactate clearance with neurodevelopmental outcomes.^14, 16, 19, 36^

However, few have investigated their relationship with early brain MRI findings, whereas it could provide an earlier way of neurological prognostication. Filling this gap is particularly important considering that most existing studies are based on small cohorts, which limits both the statistical power and the generalizability of their findings.

For instance, a 2018 prospective study involving 51 neonates with moderate to severe HIE, found that persistently elevated arterial lactate levels (measured until 36^th^ hours of life) during TH were associated with adverse motor outcomes at 24 months.^36^ Similarly a 2016 study of 17 neonates with moderate to severe HIE reported that elevated serum lactate levels after cooling and abnormal brain MRI were associated with poor neurodevelopmental outcomes at 24 months.^19^ These findings suggest a potential link between lactate level and early brain injury, but the limited sample sizes constrain the robustness of the conclusions. Other studies have specifically examined the prognostic role of lactate kinetics. For example, Heljić et al.^14^ reported that although initial lactate levels at admission were not predictive, a rapid decline in lactate during the first 24 hours of TH was associated with favorable early outcomes, including neurological exam at discharge and brain MRI findings. These results underscore the relevance of lactate trajectories over single-point measurements. In a complementary approach, O’Boyle et al.^13^ demonstrated the added value of incorporating umbilical cord blood lactate into a predictive model that combined metabolic and clinical variables. Their model improved early prediction of neurodevelopmental outcomes in neonates with HIE, supporting the integration of metabolic markers into risk stratification frameworks. More recently, Boerger et al.^16^ supported the association between elevated lactate and adverse outcomes, showing that higher initial and maximal blood lactate levels were independently associated with death or unfavorable neurodevelopmental outcome at 18-24 months of age. Although they also assessed lactate clearance, no significant association was found, potentially due to methodological limitations. Notably, they used a lower lactate normalization threshold (2 mmol/L versus 2.5 mmol/L in our study), and a substantial proportion of patients (11 out of 45) never normalized their lactate levels, possibly reducing the interpretability of clearance as a prognostic marker. Taken together, these studies point to a potential role for lactate parameters in outcome prediction, however, the evidence remains fragmented. One key limitation across the literature, beyond the small cohort sizes, is the heterogeneity in defining lactate normalization thresholds, which range from 2 to 4 mmol/L. This lack of standardization hinders cross-study comparisons and limits the ability to draw definitive conclusions.

Moreover, while previous work has mostly focused on neonates with moderate to severe HIE treated with TH, our study broadens the scope by also including mild HIE cases and neonates not treated with TH. This expanded inclusion allows for a more comprehensive analysis of lactate kinetics across the full spectrum of encephalopathy severity and treatment exposure, offering new insights into their potential role as early biomarkers of brain injury. Prognostication in neonates with mild HIE is also important, as growing evidence suggests that even these patients may experience cognitive impairment and neuropsychological difficulties at school age.^37, 38^ It can be hypothesized that TH influences lactate metabolism and clearance, potentially acting as a confounding factor. However, in our study, the lactate parameters analyzed (peak, AUC, and TLN) did not significantly differ between neonates treated with hypothermia and those who were not. Moreover, other factors known to influence lactate levels, such as glycemia, minimum pCO₂, sepsis, liver failure and use of vasoactive drugs, were comparable between the two severity groups, thereby minimizing potential bias in the interpretation of our results. Although most patients underwent echocardiography, we did not include cardiac function data in this analysis; this variable could be explored in a future study.

A major strength of our study lies in its prospective design, which minimized missing data, as well as the consistency of clinical management following a standardized care protocol for HIE. Nevertheless, several limitations should be acknowledged. First, the sample size was relatively small, although comparable to that of other neonatal studies on this topic, which may limit statistical power. Second, the predominance of mild encephalopathy in our cohort may restrict the generalizability of the findings to neonates with more severe forms of HIE. Third, the absence of a control group of healthy term neonates precludes direct comparison with baseline lactate kinetics in unaffected neonates. Finally, MR spectroscopy was not available in all patients due to technical limitations. As a result, it was not possible to assess the N-acetylaspartate and lactate peaks, which are optional components of the Weeke MRI score. However, this omission does not compromise the validity of the scoring system, as these measures are not consistently applied across cohorts in the original study.

Prognostication of outcome in neonates with HIE remains a major challenge, both in the early phase, to support decision-making regarding the initiation of TH or potential redirection of care, and in the later phase, to guide parental counseling and ensure appropriate follow-up and support for the child. Currently, no single marker, whether imaging, clinical examination, or electrophysiological assessment, provides sufficient predictive accuracy on its own. Instead, a multimodal approach is increasingly recognized as the most reliable strategy for outcome prediction. Recent studies have developed predictive models for death or neurodevelopmental impairment based on such combined modalities,^39^ while others have explored the application of machine learning algorithms to identify neonates at risk of HIE.^40^ Our study contributes to this evolving field by identifying a potential biomarker that could be integrated into future predictive frameworks.

## CONCLUSION

In this prospective cohort study of neonates with HIE, we found that blood lactate kinetics, specifically peak levels, TLN, and AUC, were significantly associated with the severity of brain injury as assessed by early MRI. These findings suggest that early lactate dynamics may serve as accessible biomarkers of cerebral injury severity in the neonatal period. Integrating these markers into clinical evaluation could enhance early prognostic accuracy and guide management strategies for a subset of this vulnerable population. Further multicenter studies are warranted to validate these results and investigate their predictive value for long-term neurodevelopmental outcomes. Further, combining functional hemodynamic data and near-infrared spectroscopy monitoring may provide further insights into the pathophysiology of lactic acid production and clearance in HIE.

## Supporting information

Supplementary Table MRI parameters

## Data Availability

All data produced in the present study are available upon reasonable request to the authors.

## ACKNOWLEDGEMENTS

This work was made possible thanks to the CIBM Center for Biomedical Imaging, founded and supported by Vaud University Hospital Centre (CHUV), University of Lausanne (UNIL), Swiss Federal Institute of Technology Lausanne (EPFL), University of Geneva (UNIGE), University Hospitals of Geneva (HUG) and the Leenaards and the Louis-Jeantet Foundations. The authors acknowledge the support of the University of Lausanne (UNIL), for covering the Open Access publication fees.

## AUTHOR CONTRIBUTIONS

G.B. contributed to the conception and design of the study, acquisition and interpretation of data, performed statistical analyses, and drafted the initial manuscript. M.D. contributed to the acquisition, analysis, and interpretation of data, supervised statistical analyses, and critically revised the manuscript. P.H. contributed to the conception and design of the study and to MRI readings, and critically revised the manuscript. J.B.L. provided input on MRI protocols, supported the clinical organization of MRI scans, and revised the radiology section of the manuscript. A.C.T contributed to the conception and design of the study and critically revised the manuscript. J.S. contributed to the conception and design of the study, interpretation of data, supervised MRI scoring, and provided substantial revisions to all sections of the manuscript. All authors approved the final version of the manuscript to be published.

## CONSENT STATEMENT

Written informed consent was obtained from the parents of all neonates included in this study.

