## Supplementary Table MRI parameters for "Blood lactate kinetics as biomarkers of MRI brain injury in neonates with hypoxic-ischemic encephalopathy"

**Supplementary Table. MRI acquisition parameters for each sequence.**

| | MP2RAGE | T2 TSE | T2 TSE | T2 EG | DTI (EPI) |
|---|---|---|---|---|---|
| **Orientation** | Sagittal | Axial | Coronal | Axial | Axial |
| **FOV (mm)** | 192x180 | 205x147 | 200x131 | 160x115 | 170x170 |
| **Matrix Size** | 192x192 | 256x256 | 256x256 | 256x210 | 100x100 |
| **Slice Thickness (mm)** | 1 | 1.6 | 1.2 | 3 | 1.7 |
| **Pixel Size ($mm^2$)** | 1 x 1 | 0.4 x 0.4 (Interpolated) | 0.4 x 0.4 (Interpolated) | 0.6 x 0.6 | 1.7 x 1.7 |
| **DF (mm)** | 0 | 0 | 0 | 0.3 | 0 |
| **Acceleration Factors** | CS x4.6 | GRAPPA 2 | GRAPPA 2 | GRAPPA 2 | SMS 4 |
| **Slices (n)** | 192 | 73 | 100 | 28 | 68 |
| **TR (ms)** | 6000 | 12000 | 5410 | 375 | 5500 |
| **TE (ms)** | 2.92 | 156 | 159 | 20 | 68 |
| **TI (ms)** | 700 / 2500 | x | x | x | x |
| **Flip Angle (deg)** | 5 | 90 | 90 | 20 | x |
| **Phase Encoding Dir.** | A > P | R > L | R > L | R > L | A > P |
| **Phase Oversampling** | 0% | 0% | 0% | 0% | 0% |
| **Number of b-value** | x | x | x | x | 2 (b = 0, 1000 s/$mm^2$) |
| **Averages** | 1 | 2 | 2 | 2 | 10 (b=0) / 1 (b=1000) |
| **Turbo/EPI Factors** | 192 | 18 | 18 | x | 100 |
| **Fat Saturation** | None | None | None | None | Yes (Strong) |
| **Bandwidth (Hz/Px)** | 240 | 235 | 219 | 200 | 1612 |
| **Excitation** | Non-sel. | x | x | Slice-sel. | Standard |
| **RF Pulse Type** | Fast | Low SAR | Low SAR | Normal | Low SAR |
| **Gradient Mode** | Fast | Whisper | Whisper | Whisper | Fast |
| **Diffusion Mode** | x | x | x | x | MDDW |
| **Directions (n)** | x | x | x | x | 20 |
| **Diffusion Scheme** | x | x | x | x | Monopolar |
| **Flow Compensation** | None | None | None | Slice/Read | x |
| **TA** | 4 min 05 s | 5 min 12 s | 4 min 58 s | 2 min 20 s | 3 min 04 s |

x = not applicable

Abbreviations**:** FOV, field of view; DF, distance factor; CS, Compressed Sensing; GRAPPA, generalized autocalibrating partially parallel acquisition; SMS, simultaneous multislice; TR, repetition time; TE, echo time; TI, inversion time; deg, degrees; A > P, anterior to posterior; R > L, right to left; b, diffusion weighting factor; Hz/Px, hertz per pixel; sel., selective; RF, radiofrequency; SAR, specific absorption rate; MDDW, multidirectional diffusion-weighted; dir., direction(s); TA, acquisition time; min., minutes; s., seconds.
